# Anti-SARS-CoV-2 vaccination does not induce the formation of autoantibodies but provides humoral immunity following heterologous and homologous vaccination regimens: Results from a clinical and prospective study within professionals of a German University Hospital

**DOI:** 10.1101/2021.11.01.21265737

**Authors:** Christoph Thurm, Annegret Reinhold, Katrin Borucki, Sascha Kahlfuss, Eugen Feist, Jens Schreiber, Dirk Reinhold, Burkhart Schraven

## Abstract

By the end of 2019 a global pandemic by the severe acute respiratory syndrome coronavirus type 2 (SARS-CoV2) causing the coronavirus disease-19 (COVID-19) has emerged. Yet, COVID-19 represents a significant economic burden to healthcare systems, destabilizes global financial markets and has caused the death of almost 5 million people worldwide. In order to prevent severe disease courses of COVID-19 especially in elderly and to establish collective immunity on the long run, different vaccines have been developed, tested and were approved within a very short time period. In Germany, the first vaccines that have been approved by local authorities were AstraZeneca’s vector virus-based vaccine Vaxzevria and the mRNA vaccines Comirnaty and Spikevax, developed by BioNTech and Moderna, respectively. As it was reported that the novel coronavirus SARS-CoV2 can trigger autoimmunity, it is of significant interest to investigate whether anti-SARS-CoV2 vaccines evoke the formation of autoantibodies and subsequent autoimmunity. Here, we did set out to systematically analyze immune responses after homologous vaccinations with mRNA or vector virus-based vaccines or after heterologous Vector/mRNA vaccinations with respect to anti-COVID-19 immune responses and, in parallel, the development of autoantibodies. In our study, we obtained serum samples one day before and 14 as well as 28 days following booster vaccination and tested them for anti-SARS-CoV2 antibodies and for autoantibodies against Cardiolipin, Prothrombin, β2-Glycoprotein, cyclic citrullinated peptides (CCP), tissue-transglutaminase (TTG) and anti-nuclear antibodies (ANA). We find that compared to homologous mRNA and heterologous Vector/mRNA vaccination, anti-SARS-CoV2 antibody levels were 90% lower after homologous vector vaccination. Of note, heterologous Vector/mRNA vaccination was found to be more effective than homologous mRNA vaccination in terms of IgM and IgA responses against SARS-CoV2. However, in terms of autoantibody generation, we only detected increases after booster vaccination in participants with already pre-existing autoantibodies. In contrast, vaccinees showing no autoantibody formation before vaccination, did not respond with sustained autoantibody production upon vaccination. Taken together, our study suggests that all used SARS-CoV2 vaccines do not significantly foster autoantibody production over time but provide humoral immunity to SARS-CoV2.

## Introduction

At the end of 2019, first cases of a novel disease causing flu-like symptoms appeared that evoked acute respiratory distress and cumulated in the death of some of the infected individuals (Harrison et al. 2020). Later, a novel and highly contagious strain of betacoronaviruses, the severe acute respiratory syndrome coronavirus type 2 (SARS-CoV2), was isolated and identified as the responsible pathogen of the disease, which was consecutively named coronavirus disease-19 (COVID-19). Since the beginning of the resulting SARS-CoV-2 pandemic, the WHO reported approximately 200 million COVID-19 cases and almost 5 million deaths as a direct consequence of COVID-19 yet (WHO 2021).

Individuals infected with SARS-CoV2 exhibit a broad spectrum of clinical symptoms. However, in approximately 50% of all cases, the infection shows an inapparent disease course, potentially favoring the undetected and uncontrolled spreading of the virus. Symptomatic SARS-CoV2 infections are characterized by symptoms like fever, fatigue, dry cough, headache, myalgia, sore throat, nausea and/or diarrhea (Rothan und Byrareddy 2020; Bai et al. 2020). In severe cases, COVID-19 is not only limited to the lungs but can also affect the skin, the kidneys, the nervous system and the hematological system (Qian et al. 2020). These COVID-19 infections often show a remarkable imbalance in the immune response, e.g. a hyperactivation of neutrophils and an excessive production of pro-inflammatory cytokines. It is of note that the latter are also frequently detected in patients suffering from autoimmune diseases (Liu et al. 2021b). Thus, from an immunological point of view, the immune reaction in response to SARS-CoV2 infections shares similarities to autoimmune diseases. In this context it was reported that COVID-19 patients carrying anti-nuclear antibodies (ANAs), anti-phospholipid antibodies or anti-SS-A/Ro show more severe courses than patients devoid of these autoantibodies (Amezcua-Guerra et al. 2021; Fujii et al. 2020; Pascolini et al. 2021; Gomes et al. 2021). Interestingly, also in non-symptomatic COVID-19 patients and patients with only mild disease courses an age-dependent tendency towards the development of autoantibodies such as anti-cyclic citrullinated peptides (CCP)-IgG and anti-tissue transglutaminase (TTG)-IgA antibodies was detected (Lingel et al. 2021).

Yet, priming of specific immune responses by active immunization represents the standard and most successful prophylactic strategy to stop the pandemic of SARS-CoV2. Therefore, different COVID-19 vaccines including mRNA vaccines, viral vectors, inactivated SARS-CoV2 or protein-based vaccines were developed (Izda et al. 2021). These efforts resulted in the approval of three particular COVID-19 vaccines by the end of 2020 in Germany. Two of them, BioNTech’s Comirnaty and Moderna’s Spikevax, use the mRNA technique to induce anti-SARS-CoV2 immunity whereas Vaxzevria developed by AstraZeneca uses a non-replicating chimpanzee adenovirus. All three vaccines appeared safe and showed significant efficacy in clinical trials upon homologous prime/boost vaccinations (Baden et al. 2021; Polack et al. 2020; Voysey et al. 2021). However, it represents a significant gap in our knowledge that despite the connection of severe COVID-19 infections and the occurrence of autoantibodies, no systematic clinical study assessing whether COVID-19 vaccination also triggers the production of autoantibodies has been published yet. Such an investigation would be of significant interest especially in view of very recent studies reporting myocarditis after mRNA vaccination especially in younger (< 30 years) male individuals (Barda et al. 2021; Mevorach et al. 2021; Verma et al. 2021; Witberg et al. 2021). To address this important question, we here analyzed anti-SARS-CoV2 and autoantibody responses in healthcare workers before and after applying heterologous and homologous vaccination protocols against COVID-19.

## Methods

### Participants

The participants of this manuscript are from the CoVac study, which was approved by the ethics board of the Medical Faculty, Otto-von-Guericke University, Magdeburg (certificate 67/21) and registered with the Paul-Ehrlich-Institute, Langen, Germany (NIS613). The CoVac study started in April 2021 and is an ongoing, prospective, observational study surveying the induction of autoimmunity upon anti-COVID-19 vaccination. Participants were recruited within the employees of the University Hospital Magdeburg and the Medical Faculty of the Otto-von-Guericke-University Magdeburg, based on prime injections and/or decision for booster injections. Participants received prime injections with Spikevax, Vaxzevria or Comirnaty. Homologous booster doses following primes by Spikevax and Comirnaty were applied 28 and 41 days, respectively. After Vaxzevria prime, participants could opt for a Spikevax or Vaxzevria boost 80 days after the prime. Participants who received a homologous vaccination with either Spikevax or Comirnaty were combined in the mRNA/mRNA group (**Table 1**). After informed consent, we collected blood samples by venipuncture followed by serum collection. Participants consisted of 36% male and 64% female with a mean age of 36 years (20-65 years) for the mRNA/mRNA group, 47 years (24-69 years) for the Vector/Vector group and 38 years (19-66 years) for the Vector/mRNA group.

**Table 1:** Basic characteristics of the study groups. Spx – Spikevax, Com - Comirnaty

| Subgroup | mRNA/mRNA n=41 |  | Vector/Vector<br>n=38 | Vector/mRNA<br>n=42 |
| --- | --- | --- | --- | --- |
|  | Spx/Spx n=25 | Com/Com n=16 |  |  |
| Age (years) | 35.9 ($\pm 13.3$) | | 47.2 ($\pm 13.5$) | 37.9 ($\pm 14.1$) |
| Sex |  |  |  |  |
| male | 16 (39%) |  | 15 (39%) | 13 (31%) |
| female | 25 (61%) |  | 23 (61%) | 29 (69%) |
| Boost (Prime + xx days) | 34.2 ($\pm 6.8$) | | 78.0 ($\pm 10.0$) | 80.3 ($\pm 5.8$) |
| 1st serum sample (Prime + xx days) | 33.2 ($\pm 6.9$) | | 77.0 ($\pm 9.7$) | 78.9 ($\pm 5.9$) |
| 2nd serum sample (Boost + xx days) | 14.5 ($\pm 1.5$) | | 14.1 ($\pm 1.2$) | 14.0 ($\pm 0.9$) |
| 3rd serum sample (Boost + xx days) | 28.2 ($\pm 1.2$) | | 28.3 ($\pm 2.5$) | 27.8 ($\pm 0.9$) |

### Quantification of Anti-SARS-CoV2-Sp1 antibodies

Quantification of anti-SARS-CoV2-Sp1-IgG antibodies was conducted using EliA SARS-CoV-2-Sp1 IgG test (ThermoFisher Scientific, Freiburg, Germany) according to manufacturer’s instructions. Samples exceeding the range of the test were diluted (1:2, 1:5 or 1:10) using sample diluent (ThermoFisher Scientific, Freiburg, Germany). To allow a better inter-assay comparison of the results, values of the EliA SARS-CoV-2-Sp1 IgG test were transposed from EliA U/mL to the BAU/mL of the WHO International Standard according to the following equation: 1 EliA U/mL = 4 BAU/mL. Quantification of anti-SARS-CoV2-IgG/IgA/IgM was performed using the Elecsys Anti-SARS-CoV-2 assay (Roche Diagnostics, Rotkreuz, Switzerland). Samples were diluted 1:20 or 1:50. The cutoffs for positive samples were ≥28 BAU/mL for the EliA und ≥0.8 BAU/mL for the Elecsys, respectively.

### Determination of neutralizing antibodies

The neutralization of the binding of the SARS-CoV2-spike protein to human ACE2 was analysed using the SARS-CoV-2-NeutraLISA (Euroimmun, Lübeck, Germany) according to the instruction by the manufacturer. Prior analysis, serum samples were diluted 1:5 in sample diluent. Samples with a relative neutralization of ≥25% were considered positive.

### ELISA

The presence of autoantibodies against cyclic citrullinated peptide (CCP, IgG; Medipan, Dahlewitz, Germany) and tissue transglutaminase (TTG, IgA; Generic Assays, Dahlewitz, Germany) were analyzed by ELISA according to the instructions of the manufacturer. Serum samples were diluted 1:100 and analyzed in duplicates. Cut-off for positivity were set at ≥ 30 U/mL for anti-CCP and ≥ 20 U/mL for anti-TTG. Quantifications of autoantibodies against Cardiolipin, Prothrombin and β2-Glycoprotein were conducted by ELISA using the Random Access Analyzer Alegria (Orgentec, Mainz, Germany) according to the manufacturer’s instructions. The following test strips were used: Anti-Cardiolipin Screen (ORG 215S), Anti-beta-2-Glycoprotein I Screen (ORG 221S), Prothrombin screen (ORG 241S). The applied cutoffs were: anti-Cardiolipin-Screen ≥ 10 U/mL, anti-β2-Glycoprotein ≥ 10 U/mL, Prothrombin screen ≥ 20 U/mL.

### Immunofluorescence

Serum samples were screened for the presence of autoantibodies against nuclear antigens (ANA) using immunofluorescence with the ANA HEp-2 plus kit (Generic Assays, Dahlewitz, Germany) according to the manufacturer’s instructions. Briefly, serum samples were initially diluted 1:80 in sample buffer. Positive samples were further diluted until 1:2560. The slides were evaluated using fluorescence microscopy by two independent observers.

### Statistics

Statistical analysis was performed using Prism8 (GraphPad, San Diego). Normal distribution of data sets was tested with Shapiro-Wilk test and significance analysis was performed using Mann-Whitney test or mixed-effects analysis with Tukey’s multiple comparison test.

## Results

### Characterization of the study cohort

Between April and August 2021, healthcare professionals of the University Hospital and the Medical Faculty of the Otto-von-Guericke-University in Magdeburg, Germany, were recruited to participate in the CoVac study. Following prime vaccination, volunteers were assigned to three individual study groups based on anti-COVID-19 boost vaccination. Participants that received a homologous vaccination with Spikevax (n=25) or Comirnaty (n=16) were pooled in the homologous mRNA group (mRNA/mRNA, n=41). The homologous vaccination group with Vaxzevria (Vector/Vector) included 38 participants whereas 42 participants underwent a heterologous vaccination regimen by first receiving Vaxzevria followed by Spikevax (Vector/mRNA) (**Table 1**). One participant from the Vector/Vector group was excluded from the study as a result of a current immunosuppressive therapy. Basic characteristics of the three groups are summarized in **Table 1**. First serum samples were obtained 33.2 days (±6.9 days), 77.0 days (±9.7 days) and 78.9 days (±5.9 days) after the prime for the mRNA/mRNA, Vector/Vector and Vector/mRNA group, respectively. Timing differences between the groups are a result of the guidelines for timing of boost vaccination following vector or mRNA vaccination provided by the German standing committee on vaccination (STIKO) of the Robert-Koch-Institute, Berlin (Vygen-Bonnet et al. 2021b).

### Both heterologous and homologous vaccination strategies provide humoral immunity to SARS-CoV2

To monitor the anti-SARS-CoV2 antibody response upon immunization, serum samples were obtained one day prior boost as well as 14 and 28 days after boost (**Figure 1A**). Samples were analyzed for the presence of anti-SARS-CoV2-Sp1-IgG antibodies. Importantly, we already detected significant differences between the study groups in the samples that were taken one day before boost. Indeed, upon vector prime, 31 samples (39%) did not show anti-Sp1-IgG antibodies above the cutoff, while following mRNA prime IgG antibodies could not be detected only in one single sample (2%) (**Figure 1B**). Further, the mean antibody concentration was 10-times lower in the vector group compared to the mRNA group (48.2 BAU/mL, 95%CI 39.4-56.9 and 527.3 BAU/mL, 95%CI 367.0-687.7, respectively).

**Figure 1:**
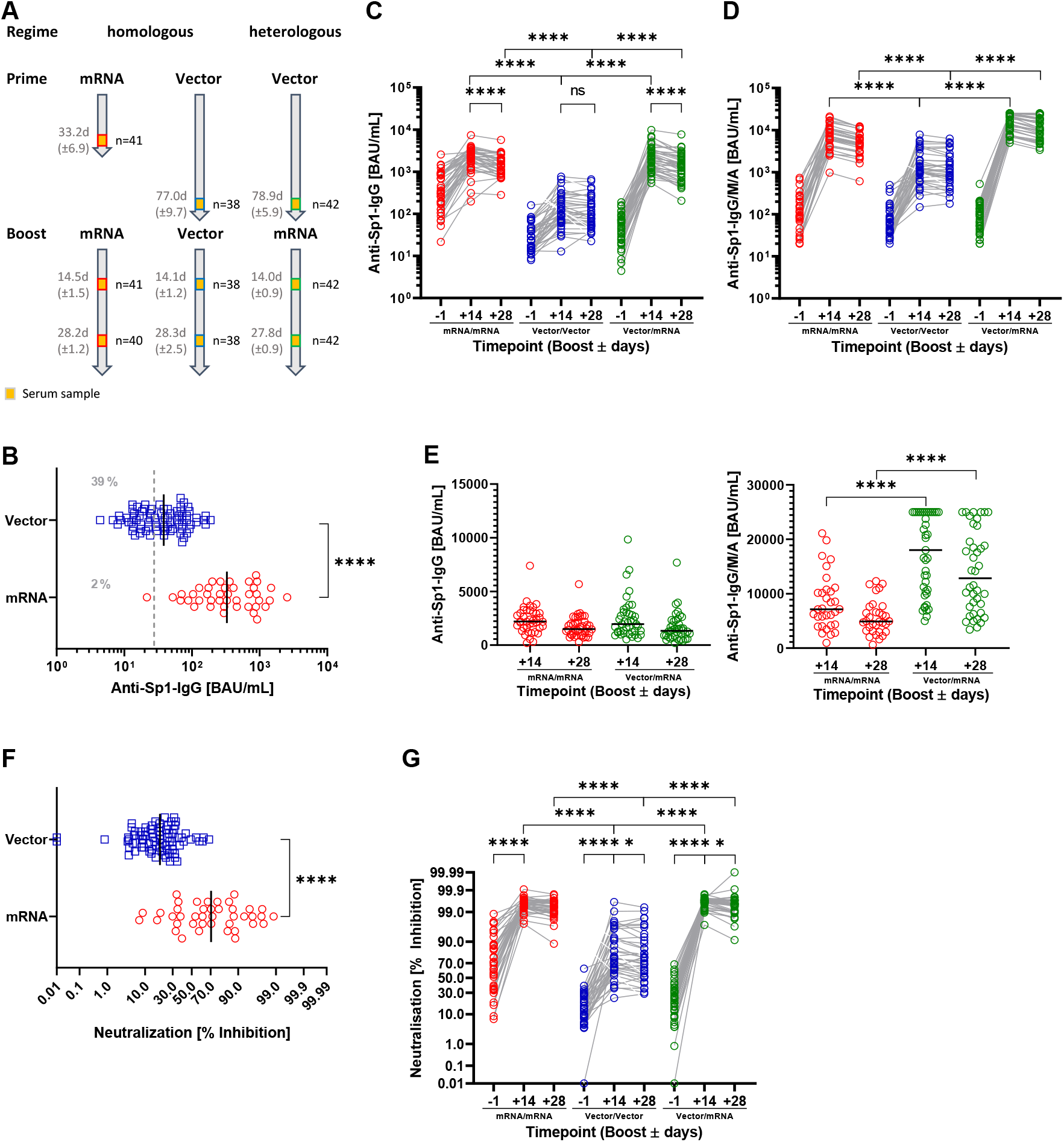
Homologous and heterologous vaccination regimens induce anti-SARS-CoV-2 antibody production and immunity. **(A)** Study design and sample generation. Based on the received vaccinations, participants were grouped into the mRNA/mRNA, Vector/Vector or Vector/mRNA group. Serum samples were obtained one day before as well as 14 and 28 days after booster. **(B)** Serum levels of anti-SARS-CoV-2-Sp1-IgG antibodies after vector (blue) or mRNA (red) prime one day before booster vaccination. The dashed line indicates the applied cutoff for positivity. **(C)** Profile of serum anti-SARS-CoV-2-Sp1-IgG antibodies of individual participants based on the different vaccination strategies (red – mRNA/mRNA, blue – Vector/Vector, green mRNA/Vector). Connected circles represent antibody levels of individual participants. **(D)** Profile of serum anti-SARS-CoV-2-Sp1-IgG/M/A antibodies of individual participants based on the different vaccination strategies and indicated groups. Connected circles represent antibody levels for individual participants. **(E)** Levels of anti-SARS-CoV2-Sp1-IgG and anti-SARS-CoV2-Sp1-IgG/M/A 14 and 28 days after homologous mRNA (red) or heterologous Vector/mRNA (green) vaccination. **(F)** Levels of neutralizing antibodies after Vector (blue) or mRNA (red) prime one day before booster vaccination. **(G)** Profile of neutralizing antibodies of individual participants within the indicated groups. Connected circles represent the antibody levels of an individual participant. Statistical analyses by Mann-Whitney test **(B and F)**, and Mixed-effects analysis with Tukey’s multiple comparison test within and between groups **(C, D, E, and G)**. **** p<0.0001

Homologous or heterologous boost vaccinations induced an increase in the Sp1-IgG antibodies in all three study groups. However, homologous vector vaccination appeared less potent with regard to antibody production compared to homologous or heterologous vaccination using an mRNA vaccine. Indeed, the mean antibody concentration in the Vector/Vector group at 14 days post boost was only 171.6 BAU/mL (95%CI 113.9-229.4) whereas it was 2,351 BAU/mL (95%CI 1,944-2,758) in the mRNA/mRNA and 2,462 BAU/mL (95%CI 1,876-3,047) in the Vector/mRNA group. No difference between the two groups after mRNA prime was detected (**Figure 1C**). We obtained similar results when we analyzed the total anti-SARS-CoV2-Sp1 antibodies including IgG, IgA and IgM. Here, mean antibody concentrations in the Vector/Vector group were also significantly lower (1,872 BAU/mL, 95%CI 1,274-2,471) compared to all other groups (Vector/mRNA 17,244 BAU/mL, 95%CI 14,930-19,557 and mRNA/mRNA 8,370 BAU/mL, 95%CI 6,534-10,206) (**Figure 1D**). Using the latter assay we detected significantly increased antibody production upon heterologous vaccination compared to homologous mRNA vaccination, which we, however, did not find when only analyzing the production of IgG antibodies, indicating that heterologous vaccination triggers stronger IgA and IgM responses (**Figure 1E**). Also at 28 days post vaccination, the antibody concentrations in the Vector/Vector group were significantly lower compared to the other two groups (**Figure 1C**) and we observed significantly increased antibody levels in the Vector/mRNA group compared to the mRNA/mRNA group when measuring anti-SARS-CoV2-Sp1-IgG/IgM/IgA (**Figure 1D** and **1E**). Of note, anti-Sp1-IgG antibody levels in the Vector/Vector group (154.4 BAU/mL, 95%CI 103.5-205.4) remained stable between day 14 and 28 post boost whereas they declined in the mRNA/mRNA (1,714 BAU/mL, 95%CI 1,404-2,025) and Vector/mRNA (1,640 BAU/mL, 95%CI 1,227-2,053) groups (**Figure 1C**).

Recent studies suggested that complete vaccination (prime and boost) using vector vaccines induce a weaker generation of neutralizing antibodies compared to mRNA vaccines (Barros-Martins et al. 2021; Hillus et al. 2021; Liu et al. 2021a; Schmidt et al. 2021; Tenbusch et al. 2021). Confirming these results, the vector group exhibited significantly reduced neutralization compared to the mRNA group in our study (21.0% inhibition, 95%CI 17.6-24.3 and 64.1% inhibition, 95%CI 54.4-73.7, respectively) (**Figure 1F**). Although we observed a significant increase in neutralization regardless of the used vaccine at 14 days post boost (**Figure 1G**), virus neutralization was again significantly lower in the Vector/Vector group compared to the mRNA/mRNA group (75.2% inhibition, 95%CI 67.9.-82.5 and 99.4% inhibition, 95%CI 99.2-99.6, respectively). Interestingly, with a heterologous booster a level of neutralization equal to the mRNA/mRNA group was achieved (99.5% inhibition, 95%CI 99.4-99.7). At 28 days post boost, neutralizations in the mRNA/mRNA and Vector/mRNA groups (98.95 inhibition, 95%CI 98.3-99.6 and 99.1% inhibition, 95%CI 98.7-99.6, respectively) were still superior compared to the Vector/Vector group (72.0% inhibition, 95%CI 64.7-79.4). Notably, we observed a significant reduction of the neutralization in the Vector/Vector and Vector/mRNA group between day 14 and day 28 whereas the mRNA/mRNA group remained stable (**Figure 1G**).

When analyzing the influence of age and gender on antibody production, we observed a slight decrease in anti Sp1-IgG levels in participants ≥50 years within the mRNA/mRNA group compared to the respective younger subgroup at 28 days post booster (**Figure S1A**). Despite being only significant at this time point, we found this trend for all time points in the mRNA/mRNA and vector/mRNA groups. In contrast, following vector boost we noted a tendency of elevated anti-Sp1-IgG antibodies in participants ≥50 years compared to younger counterparts (**Figure S1A**). We did not see any changes with respect to gender for the anti-Sp1-IgG or for the anti-Sp1-IgG/M/A as well as for neutralizing antibodies for both gender and age (**Figure 1B-F**).

### Reactogenicity

Reactogenicity was assessed one day before (for the prime) and 14 as well as 28 days after boost. To this end, participants of the study completed a questionnaire assessing local and systemic symptoms. After the prime doses, frequencies of local and/or systemic reactions were comparable between participants receiving mRNA or vector vaccination with a minority experiencing no and the majority experiencing local and systemic reactions (**Figure 2A**). However, after vector prime more participants reported only systemic reactions (e.g. fever, fatigue, chills, headache malaise or limb pain) whereas after mRNA prime more local reactions were noted. The severity of symptoms after the prime was higher in the vector group, as the majority of participants in this group (>50%) reported more than three symptoms whereas more than 70% of the vaccinees described less than three symptoms after mRNA prime (**Figure 2B**). All the reported reactions to the vaccines were mild to moderate and none severe. Compared to vector vaccination, local reactions were more frequent after prime vaccination with an mRNA vaccine as reflected by increased rates of tenderness/pain (approx. 90% after mRNA prime vs. 58% after vector prime) and erythema (approx. 22% after mRNA prime vs. 11% after vector prime) (**Figure 2C**). In contrast, vector prime vaccination induced more systemic reactions like fever (approx. 15% after mRNA prime vs. 24% after vector prime), fatigue (approx. 27% after mRNA prime vs. 50% after vector prime), chills (approx. 12% after mRNA prime vs. 39% after vector prime), headache (approx. 22% after mRNA prime vs. 49% after vector prime), malaise (approx. 10% after mRNA prime vs. 29% after vector prime) or limb pain (approx. 7% after mRNA prime vs. 28% after vector prime) (**Figure 2D**).

**Figure 2:**
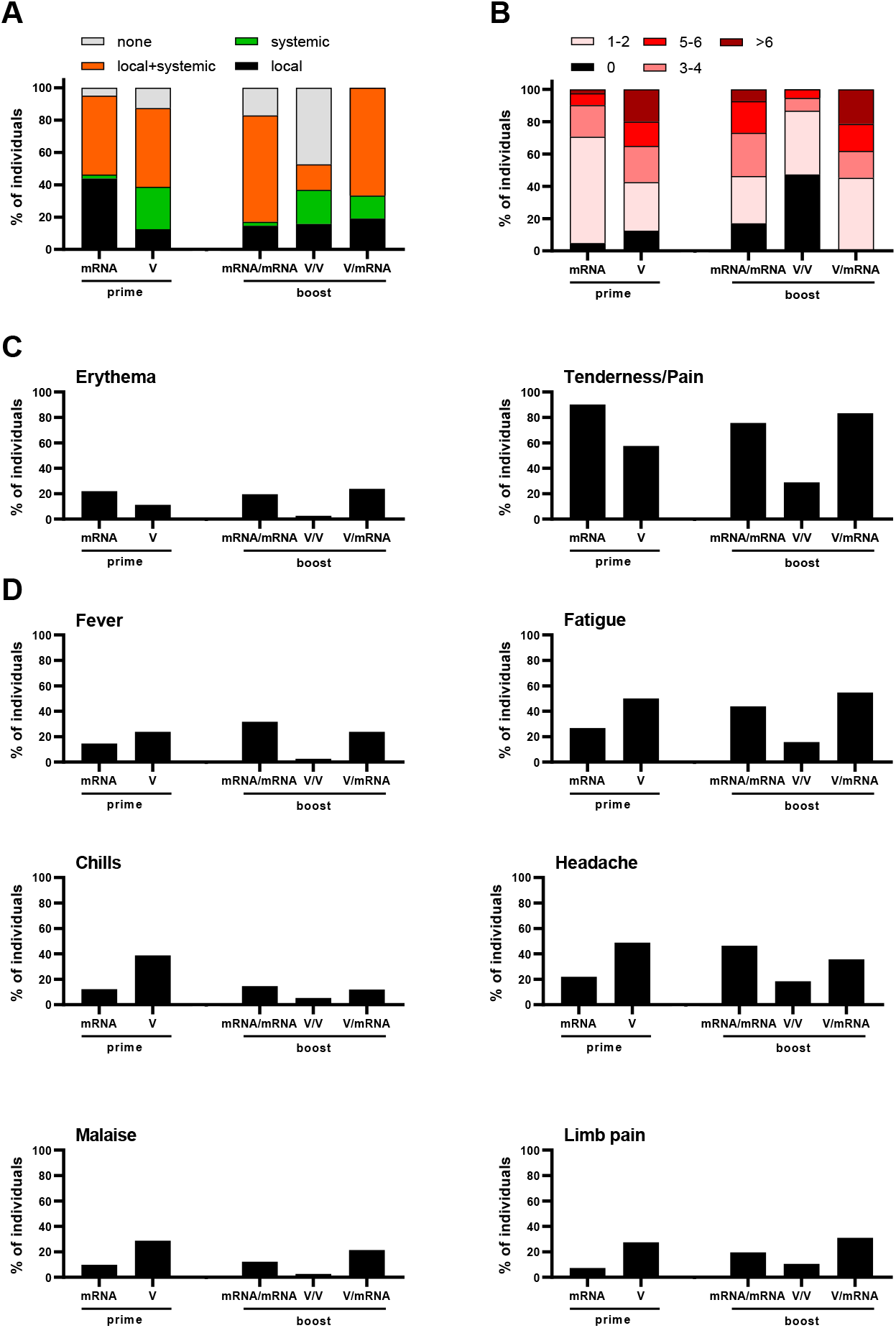
Reactogenicity after prime vaccinations with an mRNA or Vector vaccine and after booster vaccination for the mRNA/mRNA, Vector/Vector (V/V) and Vector/mRNA (V/mRNA) groups. **(A)** Analysis of local and systemic reactions. **(B)** Analysis of the severity of reactions based on the number of symptoms. **(C)** Analysis of the frequencies of local reactions. **(D)** Analysis of the frequencies of systemic reactions.

Importantly, we observed strong differences between the three study groups after boost vaccination. Thus, booster reactions after homologous vector/vector vaccination were much milder compared to the reactions after homologous mRNA/mRNA or heterologous Vector/mRNA vaccination. Of all the participants in the Vector/Vector group 47% did not experience any local or systemic reactions after the boost compared to 12% after the prime. Amongst the 53% vaccinees showing local or systemic reactions, only 12% reported more than two symptoms (compared to 58% after prime vaccination) (**Figure 2A**). In contrast, the frequency of local and systemic reactions increased after mRNA boost compared to the prime vaccinations for both groups. In addition, the severity of the reactions in the mRNA/mRNA and Vector/mRNA groups were higher compared to the Vector/Vector group and, in the case of the mRNA/mRNA group to the prime vaccination. In both groups, >50% of the participants experienced three or more symptoms (**Figure 2C**). With respect to the frequencies of individual symptoms, no major differences between the mRNA/mRNA and Vector/mRNA groups were detected (**Figure 2C-D**). In conjunction and as reported before by others (Hillus et al. 2021; Shaw et al. 2021) our data confirm that prime vaccination with an mRNA vaccine induces milder reactions compared to the vaccination with a vector vaccine. In contrast, after homologous or heterologous booster vaccination with an mRNA vaccine, the severity of reactions increased compared to homologous vaccination with a vector vaccine. However, no major differences between the mRNA/mRNA and Vector/mRNA group were observed.

### Homologous and heterologous vaccination strategies do not induce the generation of autoantibodies

In the course of infections with SARS-CoV2 but also upon vaccination, the induction of autoantibodies against various autoantigens has been described (Amezcua-Guerra et al. 2021; Fujii et al. 2020; Greinacher et al. 2021; Liu et al. 2021b). However, systematic clinical studies that investigate autoantibody production upon vaccination in a larger cohort are missing so far. Therefore, we set out to analyze whether the booster vaccination may trigger an increase in the serum concentration of distinct autoantibodies. First, we studied the profile of anti-Cardiolipin (CARD), anti-Prothrombin (PRO) and anti-β2-Glycoprotein (β2GP) autoantibodies, which are important parameters in the diagnosis of anti-phospholipid syndrome (APS) (Miyakis et al. 2006). Upon booster vaccination, two participants that had been negative after the first vaccination developed autoantibodies above the cutoff. Both individuals belonged to the Vector/Vector group. Among these, one participant showed a transient appearance of CARD autoantibodies at 14 days post booster, which declined below the cutoff at 28 days post booster (**Figure 3A**). The second participant developed PRO autoantibodies after booster vaccination, which stayed elevated at 14 and 28 days post booster (**Figure 3B**). Moreover, individuals with preexisting CARD antibodies showed a tendency of increased autoantibody concentrations throughout the observation period (**Figure 3A** and **Table 2**). This could indicate that the stimulation of the immune system by the booster vaccination with mRNA or vector vaccines also drives the production of CARD autoantibodies.

**Figure 3:**
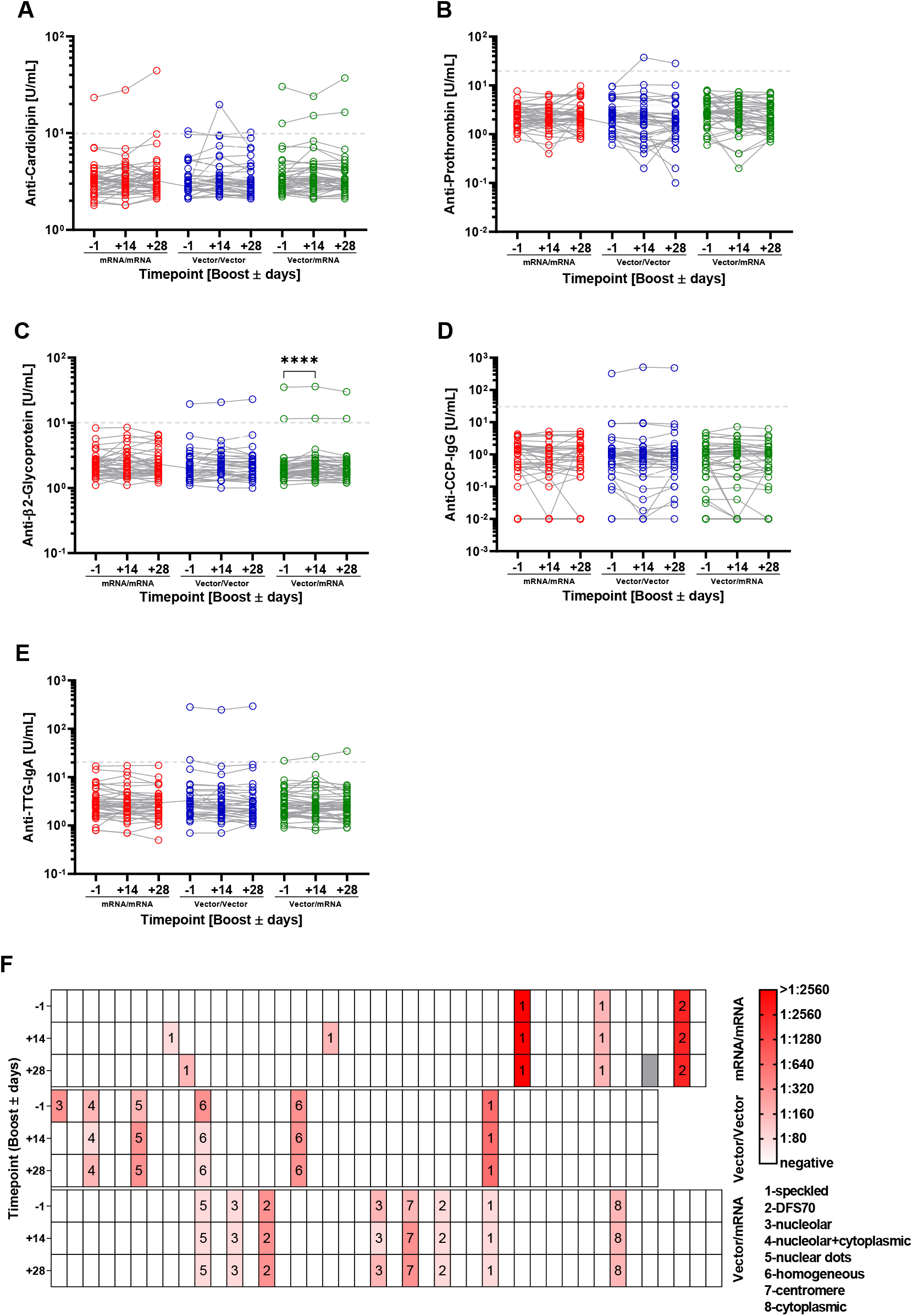
Analysis of autoantibody levels in the serum of the participants at the different time points and in the different groups (red – mRNA/mRNA, blue – Vector/Vector, green – Vector/mRNA). Connected circles represent the antibody levels of an individual participant. **(A)** anti-Cardiolipin, **(B)** anti-Prothrombin, **(C)** anti-β2-Glycoprotein,**(D)** anti-CCP,**(E)** anti-TTG autoantibody levels in the indicated groups. **(F)** Analysis of ANAs in serum samples of the shown groups. Color of the boxes indicate individual titers and numbers state the respective patterns. Statistical analyses by Mixed-effects analysis with Tukey’s multiple comparison test within and between groups **(C)**. **** p<0.0001

**Table 2:**
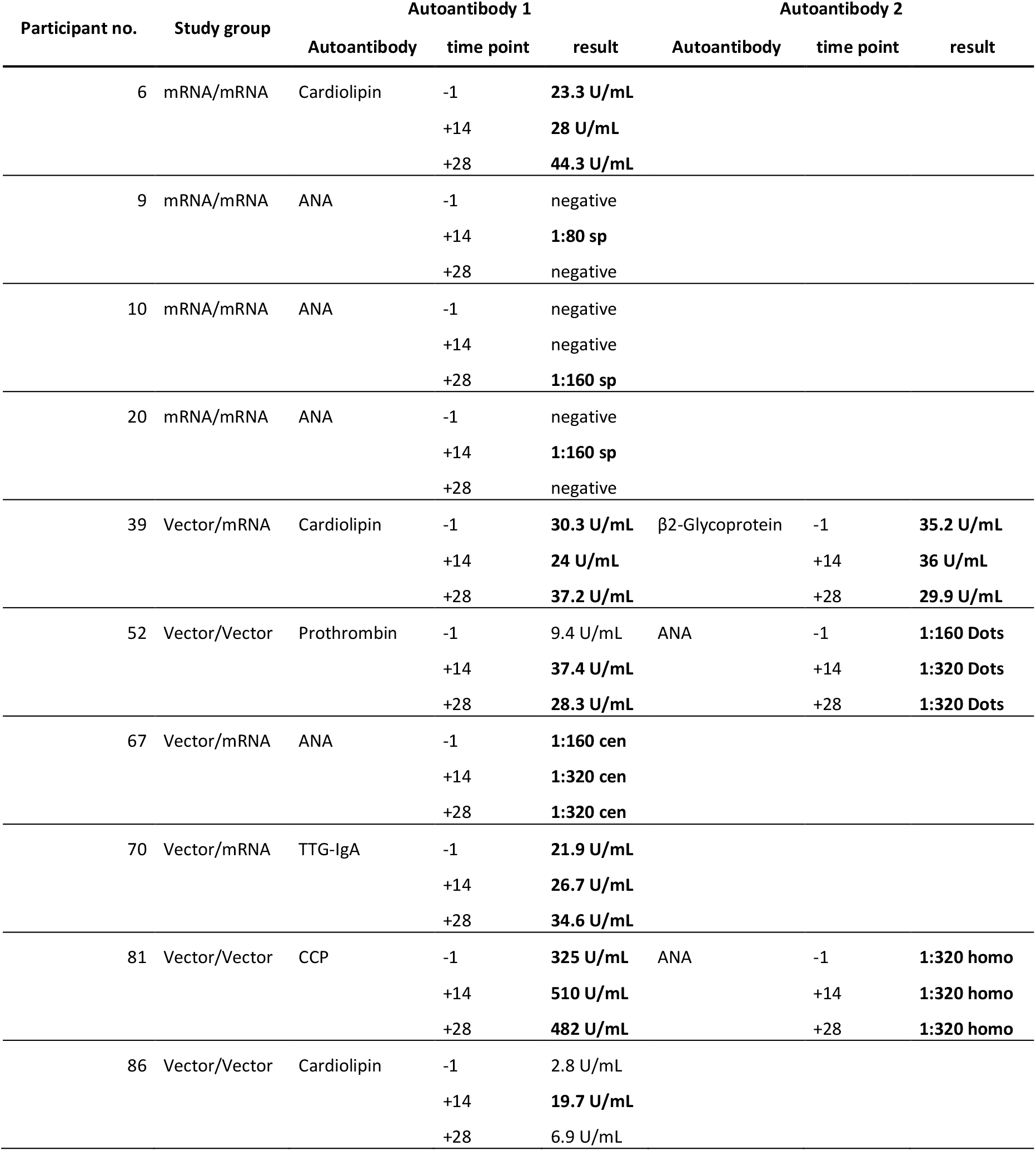
Summary of participants with increasing values for autoantibodies. Bold values are above the respective cutoff. sp – speckled, Dots – nuclear dots, cen – centromere, homo - homogeneous

| Participant no. | Study group | Autoantibody 1 |  |  | Autoantibody 2 |  |  |
| --- | --- | --- | --- | --- | --- | --- | --- |
|  |  | Autoantibody | time point | result | Autoantibody | time point | result |
| 6 | mRNA/mRNA | Cardiolipin | -1 | **23.3 U/mL** |  |  |  |
|  |  |  | +14 | **28 U/mL** |  |  |  |
|  |  |  | +28 | **44.3 U/mL** |  |  |  |
| 9 | mRNA/mRNA | ANA | -1 | negative |  |  |  |
|  |  |  | +14 | **1:80 sp** |  |  |  |
|  |  |  | +28 | negative |  |  |  |
| 10 | mRNA/mRNA | ANA | -1 | negative |  |  |  |
|  |  |  | +14 | negative |  |  |  |
|  |  |  | +28 | **1:160 sp** |  |  |  |
| 20 | mRNA/mRNA | ANA | -1 | negative |  |  |  |
|  |  |  | +14 | **1:160 sp** |  |  |  |
|  |  |  | +28 | negative |  |  |  |
| 39 | Vector/mRNA | Cardiolipin | -1 | **30.3 U/mL** | β2-Glycoprotein | -1 | **35.2 U/mL** |
|  |  |  | +14 | **24 U/mL** |  | +14 | **36 U/mL** |
|  |  |  | +28 | **37.2 U/mL** |  | +28 | **29.9 U/mL** |
| 52 | Vector/Vector | Prothrombin | -1 | 9.4 U/mL | ANA | -1 | **1:160 Dots** |
|  |  |  | +14 | **37.4 U/mL** |  | +14 | **1:320 Dots** |
|  |  |  | +28 | **28.3 U/mL** |  | +28 | **1:320 Dots** |
| 67 | Vector/mRNA | ANA | -1 | **1:160 cen** |  |  |  |
|  |  |  | +14 | **1:320 cen** |  |  |  |
|  |  |  | +28 | **1:320 cen** |  |  |  |
| 70 | Vector/mRNA | TTG-IgA | -1 | **21.9 U/mL** |  |  |  |
|  |  |  | +14 | **26.7 U/mL** |  |  |  |
|  |  |  | +28 | **34.6 U/mL** |  |  |  |
| 81 | Vector/Vector | CCP | -1 | **325 U/mL** | ANA | -1 | **1:320 homo** |
|  |  |  | +14 | **510 U/mL** |  | +14 | **1:320 homo** |
|  |  |  | +28 | **482 U/mL** |  | +28 | **1:320 homo** |
| 86 | Vector/Vector | Cardiolipin | -1 | 2.8 U/mL |  |  |  |
|  |  |  | +14 | **19.7 U/mL** |  |  |  |
|  |  |  | +28 | 6.9 U/mL |  |  |  |

We next evaluated the changes in autoantibody concentrations of the whole cohort. We did not observe any significant changes for CARD and PRO autoantibodies after the different booster vaccinations (**Figure 3A-B**). Detailed analyses of gender and age dependent effects revealed a slight decrease in CARD antibodies in individuals ≥50 years in the Vector/Vector group. With respect to PRO autoantibodies, we observed lower concentrations in individuals <50 years in the Vector/Vector group. In the Vector/mRNA group, we detected increased PRO autoantibodies in female participants compared to the respective male cohort (**Figure S2A-B**). The analysis of β2GP autoantibodies in the Vector/mRNA group revealed a slight but significant increase at 14 days post booster (**Figure 3C**). This effect was not age-dependent as individuals below and above 50 years showed the same tendency. Interestingly, this trend was exclusively observed among the female participants (**Figure S2C**). Individuals with preexisting β2GP autoantibodies did not show increasing levels within the observation period, in contrast to the CARD autoantibodies (**Figure 3C** and **Table 2**).

The analysis of the autoantibody profile in non-symptomatic and COVID-19 patients with a mild course of the disease had shown age-dependent elevations of anti-CCP-IgG and anti-TTG-IgA autoantibodies (Lingel et al. 2021). Therefore, we analyzed if the three different vaccination regimens would trigger similar events. However, we did not observe an induction in anti-CCP-IgG or anti-TTG-IgA antibodies irrespective of the vaccination regime, age or gender (**Figure 3D-C** and **Figure S2D-E**). Here, only a small fraction of the participants with preexisting autoantibodies exhibited an increase in the respective autoantibodies indicating that immune activation might augment preexisting autoantibody production.

The analysis of anti-nuclear antibodies (ANA) is an excellent tool to detect a variety of autoantibodies. We therefore determined ANAs in the different study groups. We detected ANAs in the serum of 20 participants at least at one time point. Of note, the ANA positive participants were equally distributed within the three study groups (6 participants from the mRNA/mRNA group, 6 participants from the Vector/Vector group and 8 participants from the Vector/mRNA group, respectively). **Figure 3F** summarizes the titer values and respective ANA-patterns of the samples. Amongst these, we detected an increase in ANA titers upon booster vaccination in only five participants. Three of these belonged to the mRNA/mRNA group and showed ANA positivity at only one time point with a maximal titer of 1:160 and a speckled pattern. In one participant within the Vector/Vector group, the titer increased from 1:160 before booster to 1:320 at time points 14 and 28 days post booster. In these samples, we detected nuclear dots, which were not specific for Sp100 antigen. Of note, in the same samples we also detected an increase in PRO autoantibodies. In the Vector/mRNA group, one participant exhibited a centromere-like pattern with increasing titers starting at 1:160 before the booster and reaching 1:320 post booster (**Figure 3F**).

**Table 2** summarizes the results of the autoantibody analyses for nine participants showing an increase in at least one of the tested autoantibodies upon booster vaccination. Four of these belonged to the mRNA/mRNA group. However, three participants only showed a slight induction of ANA titers up to 1:160. In a recent study, 22.5% of the tested healthy controls exhibited comparable titer values (Lingel et al. 2021). Therefore, the increases in ANA titers detected in these three individuals was possibly not triggered by the booster vaccination. When these three individuals are excluded from the evaluation, it becomes evident that all but one participant showing an increase in autoantibodies were already positive before the booster vaccination.

## Discussion

The appearance of SARS-CoV2 and the COVID-19 pandemic fueled the development of vaccines with the aim to establish specific anti-COVID-19 immunity as first line of defense. To this end, novel vaccination techniques like mRNA or vector-based vaccines were developed and some of them have shown a high efficiency and safety profile (Izda et al. 2021). By December 2020, three different vaccines had been approved for vaccination against COVID-19 in Germany. Two of these were mRNA vaccines (Comirnaty and Spikevax, respectively) while the third one was a vector-based vaccine (Vaxzevria). All three vaccines confer great protection against COVID-19. However, due to the novelty of these vaccines little is known about important question such as the dynamics of virus-specific antibody responses or a potential contribution to the deregulation of the immune system. In order to shed some light on this unexplored terrain, we systematically analyzed antibody-responses after different prime/boost regimens with respect to the intended protective immunity against COVID-19 *vs*. the unintended induction of autoimmunity.

The question of antibody production upon anti COVID-19 vaccination has been raised early after the first vaccination campaigns had started because serum antibody concentrations are widely used as correlates of protection for various other viral infections like measles, HPV, pertussis, or smallpox (Griffin 2018; Kapil und Merkel 2019; Panchanathan et al. 2006; Pinto et al. 2018). Our analyses revealed a remarkable difference in the production of SARS-CoV2 specific IgG antibodies when comparing homologous vaccination with either mRNA or vector-based vaccines. Here, the amount of antibodies produced in the Vector/Vector group reflected only 10% of that seen in the mRNA group and, further, virus neutralization appeared significantly less effective. These observations confirm previous studies (Barros-Martins et al. 2021; Hillus et al. 2021; Liu et al. 2021a; Schmidt et al. 2021; Tenbusch et al. 2021). Still, the protection against COVID-19 has been proven to be comparable in clinical trials as immunization with Vaxzevria showed an efficacy up to 90% and mRNA vaccination with Comirnaty and Spikevax exhibited 95% and 94%, respectively (Baden et al. 2021; Polack et al. 2020; Voysey et al. 2021). The discrepancy between a low production of anti-SARS-Cov2 antibodies upon Vaxzevria immunization but a protection against symptomatic COVID-19 infections that is comparable to mRNA vaccination seems to be contradictory. To solve this conundrum at least two resolutions are conceivable. First, due to our limited experience with SARS-CoV2, we simply do not know the antibody threshold required for protection against the virus. Therefore, even the low antibody levels upon vaccination with Vaxzevria might be sufficient to block viral entrance and replication. Our knowledge of influenza as a prototypical respiratory virus suggests a second explanation for the observed phenomena. Since we have a great experience with influenza vaccinations due to the annual epidemics, we know that serum antibodies are not necessarily suitable correlates of protection against infections with respiratory viruses. (Coudeville et al. 2010a; Coudeville et al. 2010b; Ohmit et al. 2011). The reasons for this are on the one hand that serum antibodies do not represent the mucosal-specific immunity that is required for effective protection against the virus and on the other hand the antigenic drift and shift of the circulating viruses which occurs after vaccination might render the vaccinations partially ineffective (Calzas und Chevalier 2019; Lim et al. 2020). Therefore, also in the case of SARS-CoV2 vaccinations, serum IgG and/or neutralizing antibodies might not be the best measure for protection. Since the cellular immunity has been shown to be essential in the course of COVID-19 infections (Diao et al. 2020; Gong et al. 2020; Grifoni et al. 2020; Le Bert et al. 2020; Sattler et al. 2020), it is likely that it also contributes to the protection after vaccination and, hence, needs to be considered as well when looking for a correlate of protection. However, first reports indicate that also T cell-mediated immunity (measured by IFNγ release) is weaker upon Vaxzevria vaccination compared to mRNA vaccination (Barros-Martins et al. 2021; Schmidt et al. 2021). But also in this regard, it has to be considered that it is currently not known how strong T cell reactions need to be in order to facilitate protection against SARS-CoV2. To further address this issue, follow-up studies analyzing the humoral and cellular immunity as well as protection from the disease months after vaccination are needed. If these studies provide evidence that cellular and humoral immunity after homologous Vaxzevria immunization are weaker, a third vaccination with an mRNA vaccine might be required for these individuals.

Due to the appearance of adverse reactions following vaccinations, Germany suspended the Vaxzevria vaccination regime in March 2021. Individuals which had already received prime vaccinations with Vaxzevria could opt for a heterologous boost with an mRNA vaccine (Vygen-Bonnet et al. 2021a). In our study, this heterologous vaccination strategy was as effective in the induction of anti-Sp1-IgG antibodies as a homologous mRNA vaccination (**Figure 1C**) despite the fact that the antibody level before boost as well as the interval between the prime and the boost and were different. These observations are in line with other reports (Barros-Martins et al. 2021; Schmidt et al. 2021; Tenbusch et al. 2021). However, unlike the IgG-response, the pan-antibody response (also including IgA and IgM) is much stronger upon heterologous vaccination (**Figure 1D**). To our knowledge, this is the first report showing these strong differences. Especially with respect to the production of SARS-CoV2-specific IgA our observations might be of major interest. An important role of IgA in the protection against COVID-19 was suggested due to a correlation between the prevalence of selective IgA deficiency (sIgAD) and the occurrence of COVID-19 infections. Countries with a low prevalence of sIgAD show less COVID-19 infections (Naito et al. 2020). This observation is not without precedent as it has previously been shown that IgA Abs with neutralizing capacity against influenza virus are produced upon consecutive vaccinations against the same influenza strain (Abreu et al. 2020). Since in some western countries third COVID-19 vaccinations have started, it would be of interest to learn if the third vaccination might trigger even stronger IgA responses (and therefore improve anti-COVID-19 protection) after a heterologous prime/boost strategy. Despite the fact that the instantaneous IgA-mediated protection against respiratory viruses is mediated via soluble IgA in the mucosa, also serum IgA is able to confer immunity (Seibert et al. 2013). However, a final prediction of the importance of IgA is complicated by the observations that, unlike influenza, high IgA levels upon COVID-19 infection appear to be associated with a more severe course of the disease (Hasan Ali et al. 2020). Yet, it is more likely that severe COVID-19 induces strong IgA production rather than high pre-infection IgA levels are responsible for the severe course of COVID-19. Long-term studies are required to clarify which vaccination strategy exhibits the best protection.

In the context of COVID-19 infections, various reports described the appearance of autoantibodies like ANA, anti-phospholipid antibodies or SS-A. Furthermore, the appearance of these autoantibodies correlated with a more severe course of the disease (Amezcua-Guerra et al. 2021; Fujii et al. 2020; Pascolini et al. 2021). In order to identify the molecular and cellular basis underlying these observations, mechanisms that are known to induce autoimmunity upon viral infections, e.g. molecular mimicry, have been assessed for COVID-19. Many hexa-and heptapeptides originating from human chaperones, olfactory receptors or different membrane proteins are also part of the SARS-CoV2 proteome (Angileri et al. 2020; Kanduc und Shoenfeld 2020; Lucchese und Flöel 2020; Marino Gammazza et al. 2020). However, the biological relevance of these structures has not been proven so far. Additionally, many case reports and small studies have tried to establish a connection between COVID-19 and autoimmunity (Bowles et al. 2020; Helms et al. 2020; Shah et al. 2020; Zhang et al. 2020). However, none of the so far existing studies clearly showed that COVID-19 indeed augments autoimmune diseases and systematical studies investigating autoantibodies following different vaccination regimens are elusive yet. It is important to note that a coincidence of events does not necessarily imply causality, which means that in some cases COVID-19 and autoimmunity could represent completely unrelated events that by chance appear in parallel. Alternatively, also a pre-existing subclinical autoimmunity might contribute to these observations. Nevertheless, the discussion about a possible interconnection between autoimmunity and COVID-19 triggered the fear that vaccination might induce autoimmunity (Ehrenfeld et al. 2020). This is further emphasized by recent reports about different cases of Myocarditis upon vaccinations with mRNA vaccines (Witberg et al. 2021; Verma et al. 2021; Mevorach et al. 2021; Barda et al. 2021). To our knowledge, our study is the first that systematically addresses this issue. Importantly, we did not detect an induction of autoantibodies upon booster vaccination in all three study groups. Only β2-Glycoprotein autoantibodies slightly increased in females upon heterologous vaccination but, most notably, they remained below the cutoff. Moreover, only participants with pre-existing autoantibodies responded with detectable increases in autoantibody production upon vaccination, a phenomenon that has been described for other vaccinations before (Soriano et al. 2015). However, one participant with positive ANA result before booster vaccination developed anti-Prothrombin antibodies thereafter. Follow up analyses showed a decline in anti-Prothrombin antibodies back to baseline after four months (data not shown). Nevertheless, none of the participants had a history of autoimmunity or developed clinical signs that would indicate the occurrence of an autoimmune disease after the booster. Therefore, it is questionable whether the autoantibodies that we detected during our study are of medical relevance. Furthermore, APS antibodies can also be induced by various triggers, for example viral infections (Cruz-Tapias et al. 2012). Therefore, it cannot be ruled out that vaccination-unrelated factors/events might be responsible for the observed changes in autoantibody levels. Still, it is important to note that the induced autoantibodies either declined to baseline levels or showed no further increase during a follow up observation period (data not shown). This might indicate that the appearance of autoantibodies is a transient event that occasionally accompanies the vaccination but does not have clinical relevance. However, we certainly cannot exclude the possibility that more severe and possibly clinically relevant autoimmunity is triggered after repetitive vaccinations with the same vaccines or following vaccination with new vaccines that are developed in the future with the aim to fight novel COVID-19-mutants. We are aware that the explanatory power of this study is limited due to the relatively small group sizes and the short observation period. Hence, to clarify the question whether SARS-CoV2 vaccination can cause autoimmunity future analyses of greater cohorts appear to be mandatory. These studies should also include a healthy control group and predisposed individuals in order to improve the predictive power.

## Data Availability

All data produced in the present study are available upon reasonable request to the authors.

---

**Figure S1:**
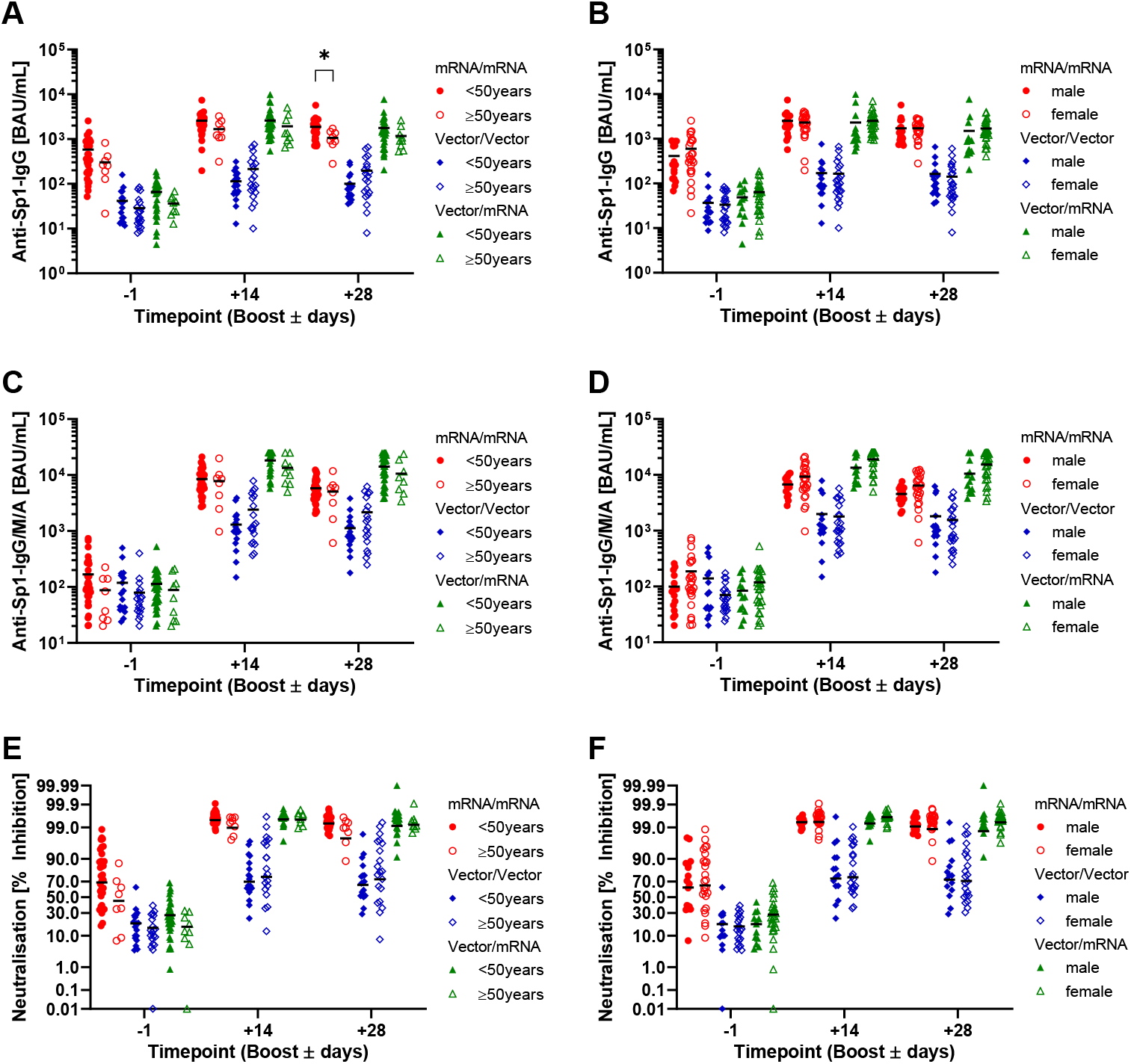
Analysis of anti-SARS-CoV2-Sp1 antibodies in the different groups (red – mRNA/mRNA, blue – Vector/Vector, green mRNA/Vector) with respect to age and gender. **(A, C, and E)** Anti-SARS-CoV2-Sp1-IgG (A), anti-SARS-CoV2-Sp1-IgG/M/A (C) and neutralizing antibodies (E) in individuals <50 years (filled symbols) and ≥50 years (open symbols). **(B, D, and F)** Anti-SARS-CoV2-Sp1-IgG (B), anti-SARS-CoV2-Sp1-IgG/M/A (D) and neutralizing antibodies (F) in males (filled symbols) and females (open symbols). Statistical analyses by Mixed-effects analysis with Tukey’s multiple comparison test **(A)**. * p<0.05

**Figure S2:**
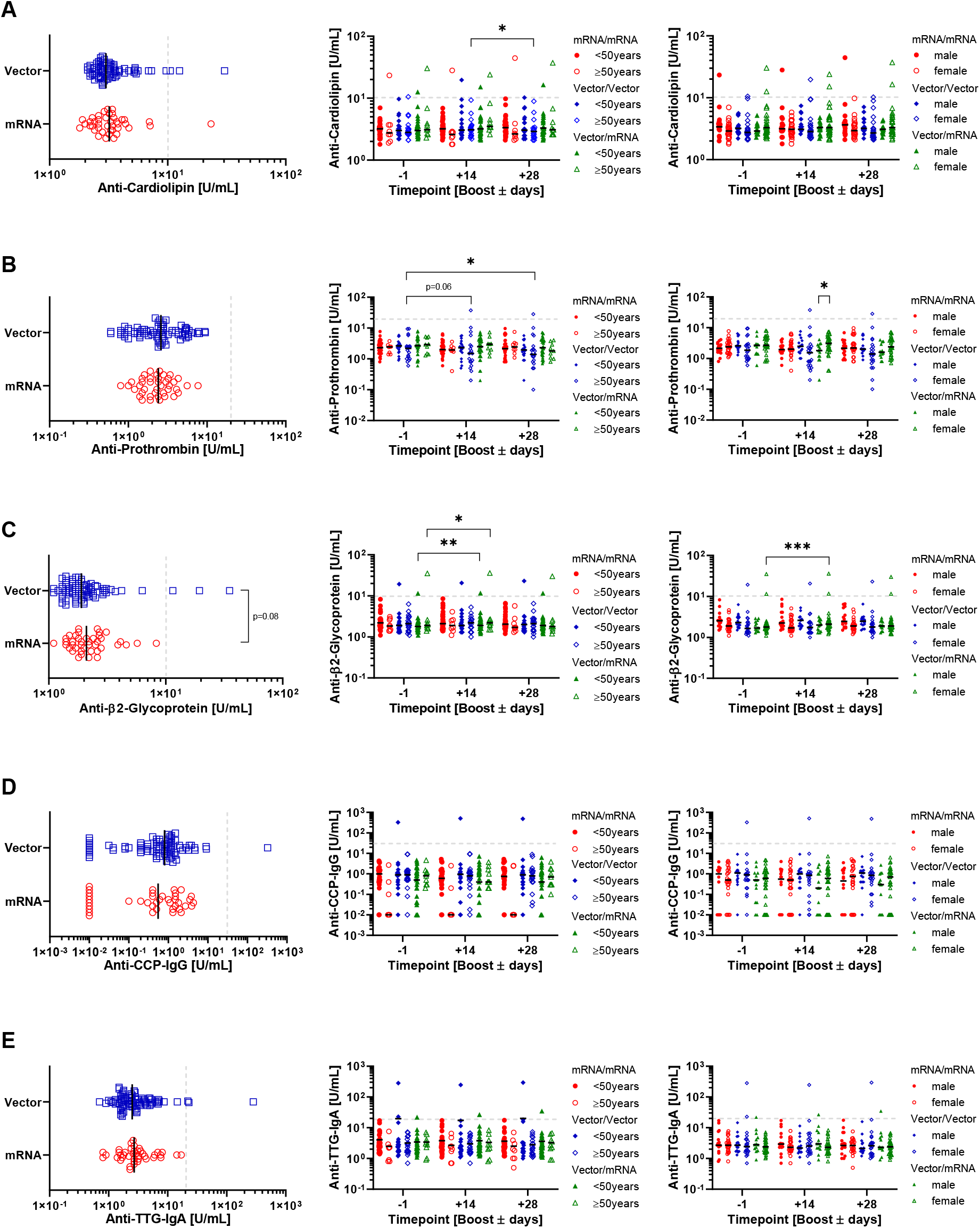
Analysis of autoantibodies based on prime vaccinations in serum samples 1 day before boost (blue – vector prime, red – mRNA prime) and in the different groups (red – mRNA/mRNA, blue – Vector/Vector, green mRNA/Vector) with respect to age and gender. **(A)** Anti-Cardiolipin antibodies: left, autoantibody levels before boost based on prime vaccinations; middle, analysis in individuals <50 years (filled symbols) and ≥50 years (open symbols) based on the study groups; right, analysis in males (filled symbols) and females (open symbols) based on the study groups. **(B)** Anti-Prothrombin antibodies: left, autoantibody levels before boost based on prime vaccinations; middle, analysis in individuals <50 years (filled symbols) and ≥50 years (open symbols) based on the study groups; right, analysis in males (filled symbols) and females (open symbols) based on the study groups. **(C)** Anti-β2-Glycoprotein antibodies: left, autoantibody levels before boost based on prime vaccinations; middle, analysis in individuals <50 years (filled symbols) and ≥50 years (open symbols) based on the study groups; right, analysis in males (filled symbols) and females (open symbols) based on the study groups. **(D)** Anti-CCP antibodies: left, autoantibody levels before boost based on prime vaccinations; middle, analysis in individuals <50 years (filled symbols) and ≥50 years (open symbols) based on the study groups; right, analysis in males (filled symbols) and females (open symbols) based on the study groups. **(E)** Anti-TTG antibodies: left, autoantibody levels before boost based on prime vaccinations; middle, analysis in individuals <50 years (filled symbols) and ≥50 years (open symbols) based on the study groups; right, analysis in males (filled symbols) and females (open symbols) based on the study groups. Statistical analyses by Mixed-effects analysis with Tukey’s multiple comparison test within and between groups **(A-C)**. * p<0.05, ** p<0.01, *** p<0,001

